# Development and evaluation of a new IgM/IgG rapid diagnostic test for SARS-CoV-2

**DOI:** 10.1101/2020.10.09.20209866

**Authors:** P.J. Ducrest

**Affiliations:** GaDia SA, Route de l’ile au bois 1A, 1870 Monthey, Switzerland

**Keywords:** SARS-CoV-2, COVID-19, serology, rapid test, Lateral Flow immunoassay, Point-of Care Testing

## Abstract

There is an urgent need in rapid diagnostic test (RDT) to detect antibodies against SARS-CoV-2. We have developed a rapid and simple point-of-care lateral flow immunoassay (LFIA) detecting IgM and IgG against SARS-CoV-2 in 10 minutes. The aim of this study is to evaluate the diagnostic performance of this RDT. RT-PCR positive plasma samples (n=35) for SARS-CoV-2 and 97 negative control samples were studied. Diagnostic performance of IgG/IgM RDT was assessed using both gold standard RT-PCR and Electro-chemiluminescence immunoassay (ECLIA) Elecsys^®^ Anti-SARS-CoV-2 total Ig. Overall, RDT sensitivity was 100% (95% confidence interval [95%CI]: 88-100%) and specificity 93% (95% CI: 85-97%). This IgG/IgM RDT done in plasma displays a high diagnostic accuracy for SARS-CoV-2 IgG/IgM in high COVID-19 prevalence settings. Its use could be considered in the absence of routine diagnostic serology facilities for samples collected between 10 and 180 days after symptoms onset.

## Introduction

SARS-CoV-2 is the etiological agent of a several cases of pneumonia, first reported in Wuhan (Hubei, China), called 2019 Coronavirus Disease (COVID-19). Protein sequence analysis of seven proteins has showed that the virus belongs to the species of Severe Acute Respiratory Syndrome Coronavirus (SARS-CoV) and Middle East Respiratory Syndrome Coronavirus (MERS-CoV) with similar epidemiology^1,2^. Currently, the detection method for SARS-CoV-2 is based on viral RNA detection using Reverse Transcribed Real-Time PCR (RT-PCR) but other tests such as chest computed tomography (CT) imaging or rapid antigen/antibody testing could be used^3^. Testing of specific antibodies of 2019-SARS-CoV-2 in patient blood is a good choice for rapid, simple, highly sensitive diagnosis of COVID-19. Moreover, Serological analysis indicates the exposure to the virus and if a patient has developed an immunity against the virus. It is widely accepted that IgM provides the first line of defense during viral infections, prior to the generation of adaptive, high affinity IgG responses that are important for long term immunity and immunological memory^3^. Several authors have analyzed the antibody kinetic in COVID-19 patients. Zhao et al. 2020, have shown that among 173 patients, the seroconversion rate for total antibody, IgM and IgG was 93.1% (161/173), 82.7% (143/173) and 64.7% (112/173), respectively^4^. The seroconversion sequentially appeared for total antibody, IgM and then IgG, with a median time of 11, 12 and 14 days after onset. The majority of antibodies are produced against the most abundant protein of the virus, which is the Nucleocapsid protein (NP). Therefore, tests that detect antibodies to NP would be the most sensitive. However, the receptor-binding domain of Spike protein (RBD-S) is the host attachment protein, and antibodies to RBD-S would be more specific and are expected to be neutralizing. Therefore, according to authors, using one or both antigens would result in high sensitivity^5^. Moreover, the source of protein (prokaryotic vs eukaryotic) as well as the region of the protein (RBD, full-length) and the tag used (His tag or Fc Tag) is a major concern in the diagnostic performance^6^.

The aim of this study is to develop and evaluate the performance of a new IgM/IgG rapid diagnostic test (RDT) based on lateral flow assay (LFA) technology, in a high COVID-19 prevalence setting using gold standard RT-PCR as well as an Electro-chemiluminescence immunoassay (ECLIA) Elecsys® Anti-SARS-CoV-2 total Ig (Roche, Switzerland).

## Material & Methods

### Preparation of rapid test IgM/IgG anti-SARS-CoV-2

Anti-human IgM was purchased from Hybridoma Reagent Laboratory Inc. (Baltimore, USA) and protein G from ThermoFisher Scientific Inc. (USA), COVID-19 recombinant Receptor Binding Domain antigen was purchased from GenScript Biotech (Netherlands) B.V. (Leiden, NL). Several different designs of antigen were tested and optimized. Bovine serum albumin (BSA) and sucrose were purchased from Sigma, BSA-biotinylated from Vector Laboratories Inc. (Burlingame, USA) and purified streptavidin from BioLegend Inc. (San Diego, USA). 80 nm gold nanoparticle (AuNP) colloids was purchased from nanoComposix Inc. (San Diego, USA), nitrocellulose membrane backed on plastic support were obtained from Merck Millipore (Darmstadt, Germany), the glass fiber conjugate pad, sample pad and absorbent pad were obtained from Ahlstrom-Munksjö (Helsinki, Finland). To prepare the AuNP conjugate, SARS-CoV-2 recombinant protein was added to AuNP colloid according to manufacturer protocol. Similarly, the control line AuNP colloid was produced using Streptavidin. The test strip consists of four parts, including nitrocellulose backed on plastic support, sample pad, conjugate pad and absorbent pad. The anti-human IgM, protein G and BSA-biotin were immobilized at test line M, G and control line (C line), respectively. Conjugate pad was sprayed with mixture of AuNP-COVID-19 recombinant antigen conjugate and AuNP-streptavidin. Test strip were then placed in a plastic housing cassette and sealed in aluminum pouch with a desiccant (figure 1). The test has been developed and produced by GaDia SA (Monthey, Switzerland).

**Figure 1:**
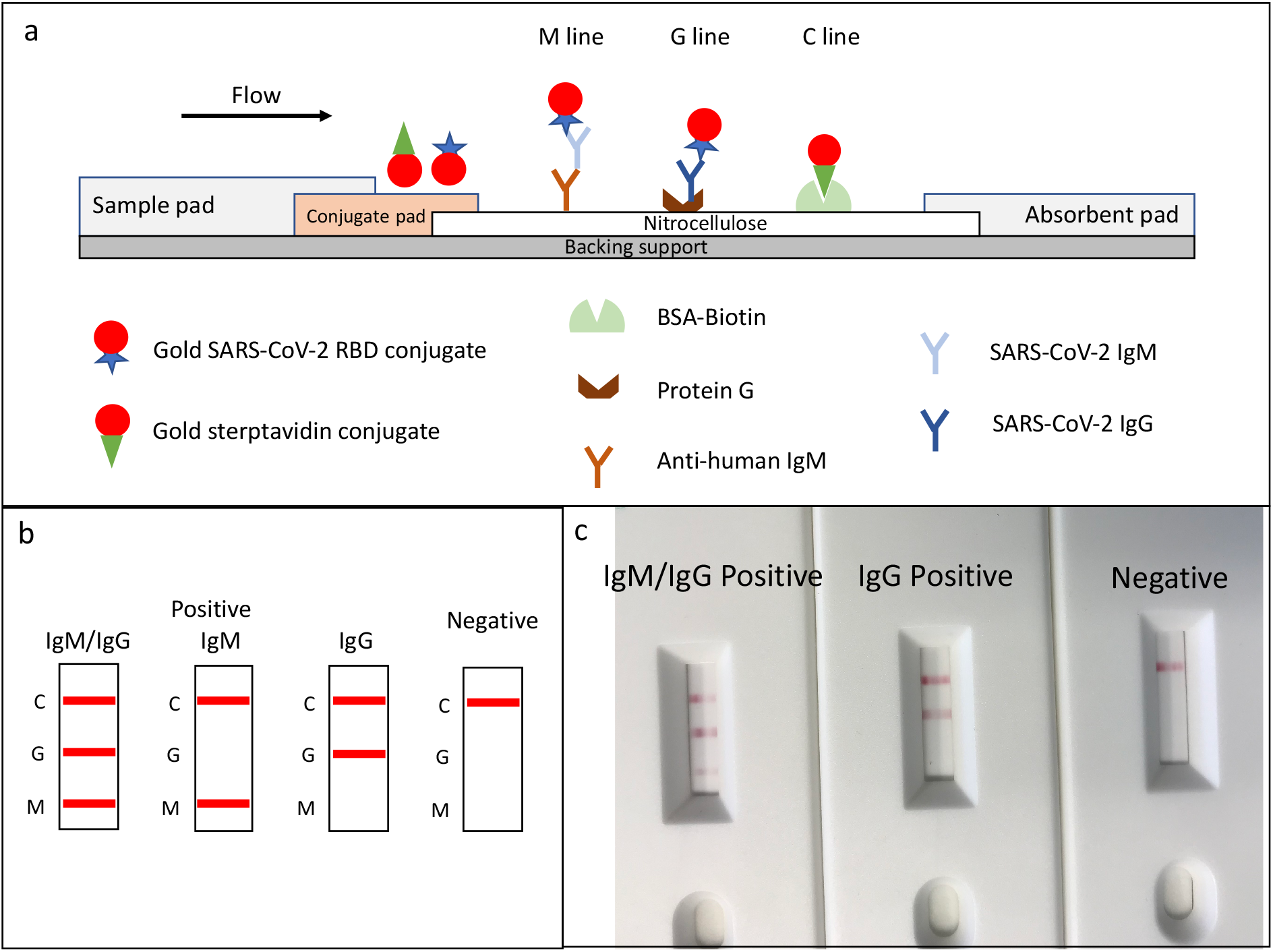
Schematic illustration of rapid SARS-CoV-2 IgM-IgG combined antibody test and example of typical results. a, Schematic diagram of the detection device; b, illustration of different testing results; c, example of typical results obtained with the RDT.

### Sample Testing

The pouched test was opened immediately before use. To perform the test, 10 µL of plasma sample was pipetted into the sample area followed by adding 2-3 drops (60-70 µL) of running buffer (PBS + 1% tween 20 + 1% BSA). The result is interpreted after 10 minutes. Three detection lines are on the stip. The control (C) line appears when sample has flowed through the strip. The presence of anti-SARS-CoV-2 IgM and/or anti-SARS-CoV-2 IgG will be indicated by a red line in the M and/or G region. If only the control line (C) appear, the sample does not contain IgM nor IgG antibodies. If the control line does not appear, the test is invalid, and the test should be repeated (figure 1).

### Study population and blood sample collection

Anonymized leftovers of diagnostic specimen plasma-EDTA, supplied by INO Specimens BioBank, ISB (Clermont-Ferrand, France), were used for the evaluation. All 35 positive samples used have a day post symptom (DPS) of at least 10 days and were positive for SARS-CoV-2 using RT-PCR (BD SARS-CoV-2 reagent kit for the BD Max system; Becton Dickinson and Co, US). Additional serological analysis was used for 11 out of 35 RT-PCR positive samples using Elecsys® Anti-SARS-CoV-2 total Ig (Roche, Switzerland). All analyses were conducted by INO Specimens BioBank. Unmatched healthy plasma samples (n=97), supplied by AbBaltis (Kent, UK) with a collection date before 2018, were used as negative control group.

### Electro-chemiluminescence immunoassay Elecsys® Anti-SARS-CoV-2 total Ig

Elecsys® Anti-SARS-CoV-2 total Ig (Roche, Switzerland) uses the nucleocapsid protein of SARS-CoV-2 as antigen. Total Ig was analyzed according to the manufacturer’s instructions. The assays were run on Cobas e 601 (Roche, Switzerland) according to the manufacturer’s protocol. Positivity is defined by manufacturer as cut-off index (COI) ≥ 1.0.

### Statistics

Vassarstats online tool (www.vassarstats.net) was used to calculate sensitivity (SE), specificity (SP), positive and negative predictive values (PPV, NPV), 95% confidence intervals, median, and Interquartile range (IQR); while significance (p-values) was calculated using Mann-Whitney U test for categorical variables. Statistical significance was defined as p < 0.05.

## Results

### Rapid test development

The SARS-CoV-2 rapid IgM/IgG antibody test is a lateral flow qualitative immunoassay for the rapid determination of the presence or absence of both anti-SARS-CoV-2-IgM and anti-SARS-CoV-2-IgG in human specimens. The test strip has three detection lines, including a control line that appears when the sample has flowed to the end of the strip. The presence of SARS-CoV-2 IgG and IgM antibodies are indicated by a red line in the specific region indicated on the device (M line or G line). When IgM or IgG are present in the sample, they bind to SARS-CoV-2 Receptor Binding Domain Antigen coated on gold Nanoparticles. Then this complex antibody-antigen-gold Nanoparticles flows through the device. The anti-SARS-CoV-2 IgM antibodies will bind to the M (IgM) line, and the IgG antibodies will bind to the G (IgG) line. If the specimen does not contain SARS-CoV-2 antibodies, no labeled complexes bind at the test zone and no M or G lines are observed. The remaining colloidal gold flows up to the control line, where streptavidin gold nanoparticles will bind to BSA-biotin. A red line must appear at the control line zone for all valid tests whether the sample is positive or negative for SARS-CoV-2 antibody (figure 1).

### Baseline characteristics

The baseline demographic characteristics of patients samples were as follows: the 35 RT-PCR confirmed COVID-19 samples from patients were older (median 50 years old, IQR 34.5-60.5) compared to the healthy plasma samples patients (n=97) (median 33.5 years old, IQR 19-43; p<0.0001). The proportion of females in COVID-19 cohort was 54% (n=19) and 56% (n=54) in healthy control group Among COVID-19 samples, the median delay between symptoms onset and sampling was 21 days (IQR 16-32 days), but not less than 10 days. The longest DPS for one single sample was 180 days. All RT-PCR tests were done in nasopharyngeal secretions. The median CT value was 25.6 (IQR 20.05-29.15). Electro-chemiluminescence immunoassay (ECLIA), Elecsys® Anti-SARS-CoV-2 total Ig (Roche, Switzerland) was conducted on 11 out of 35 RT-PCR confirmed samples. The median cut-off index (COI) was 49.1 (IQR 7.5-103.2 COI).

### Specificity of IgG/IgM RDT on negative control group

IgG/IgM RDT diagnostic specificity against negative control group with a sampling date before 2018 (n=97) is shown in Table 1. IgG/IgM RDT revealed concordant results in 90 of 97 samples. Seven discordant results showed positive results for IgM line only, while being negative (collection date before the beginning of COVID-19 outbreak) (false-positives). These resulted in an IgG/IgM RDT specificity (SP) of 93% (95% CI: 85-97%). When considering only IgG line results, the specificity was 100% (95% CI: 95-100%).

**Table 1:** Diagnostic performance of RDT IgM/IgG anti-SARS-CoV-2

|  | SE %<br>(95%CI) | SP %<br>(95%CI) | PPV %<br>(95%CI) | NPV %<br>(95%CI) |
| --- | --- | --- | --- | --- |
| **IgG/IgM RDT vs RT-PCR**<br>**(n=132), prevalence 35/132=27%** | 100 (88-100) | 93 (85-97) | 83 (68-92) | 100 (95-100) |
| **IgG/IgM RDT vs ECLIA**<br>**(n=108), prevalence 11/108=10%** | 100 (68-100) | 93 (85-97) | 61 (36-82) | 100 (95-100) |

RDT: rapid diagnostic test; SE: sensibility; SP: specificity; PPV: positive predictive value; NPV: negative predictive value; ECLIA: Electro-chemiluminescence immunoassay; CI: confidence interval

### Sensitivity of IgG/IgM RDT against RT-PCR confirmed COVID-19 samples

IgG/IgM RDT diagnostic sensitivity against RT-PCR confirmed samples (n=35) is shown in Table 1. All RT-PCR confirmed samples were positive either for IgG or IgM or both. These resulted in an IgG/IgM RDT sensitivity (SE) of 100% (95% CI: 88-100%), while the PPV was 83% (95% CI: 68-92%) and the NPV 100% (95% CI: 95-100), in this unmatched case-control study, compared to RT-PCR confirmed samples. Two samples with days post symptoms (DPS) of respectively 170 and 180 days were still positive for IgG and/or IgM (Supl. Material).

### Sensitivity of IgG/IgM RDT against Electro-chemiluminescence immunoassay

IgG/IgM RDT accuracy against Electro-chemiluminescence immunoassay (ECLIA), Elecsys® Anti-SARS-CoV-2 total Ig (Roche, Switzerland) (n=23) is shown in Table 1. Both methods revealed similar results in all samples. These resulted in an IgG/IgM RDT sensitivity (SE) of 100% (95% CI: 68-100%), while the PPV was 61% (95% CI: 36-82%) and the NPV 100% (95% CI: 95-100).

The table 2 compares the results obtained with total Ig ECLIA and the developed rapid test (RDT). A positive ECLIA result is characterized by a Cut-off index value (COI) of > 1.10. The analytical sensitivity of the rapid test is similar to the ECLIA, with low positive samples (COI = 1.19) the rapid test is positive for IgM only, indicating a probable early infection (DPS = 10).

**Table 2:** Comparison of results obtained with ECLIA Total Ig (Cobas, Roche, Switzerland) and RDT (GaDia)

| Sample # | DPS [days] | GaDia RDT |  | Cobas (Roche) |
| --- | --- | --- | --- | --- |
|  |  | IgM | IgG | Total Ig, COI [-] |
| 25 | 39 | pos | pos | 180,9 |
| 26 | 22 | pos | pos | 169,6 |
| 27 | 20 | neg | pos | 114,9 |
| 28 | 14 | pos | pos | 91,45 |
| 29 | 41 | pos | pos | 76,85 |
| 30 | 16 | pos | pos | 49,05 |
| 31 | 17 | pos | pos | 20,16 |
| 32 | 24 | pos | pos | 10,1 |
| 33 | 15 | pos | pos | 4,98 |
| 34 | 26 | pos | pos | 2,06 |
| 35 | 10 | pos | neg | 1,19 |

## Discussion

The key finding of the present evaluation study, using an unmatched case-control design including 73.5% of negative control samples, is that the diagnostic accuracy of IgG/IgM RDT on plasma samples when compared to RT-PCR or ECLIA is the same and displayed a SE of 100%, a SP of 93% and a NPV of 100%. When compared to ECLIA, positive predictive value is of 61% and 83% when compared to RT-PCR due to prevalence.

No false-negatives result was obtained with this RDT, compared to ECLIA values, indicating that even with borderline samples (Cut-of index of 1.19), the test is positive. ECLIA Manufacturer defined positive samples when Cut-of index (COI) value is ≥ 1.0. This finding indicates that the analytical sensitivity of this newly developed RDT is similar than the ECLIA Elecsys® Anti-SARS-CoV-2 total Ig (Roche, Switzerland), even if the ECLIA is targeting the immune response against full-length N protein of SARS-CoV-2 and our RDT the immune response against RBD of spike protein.

Such performances indicate that this RDT could be fit for purpose in clinical settings where a high prevalence of COVID-19 prevails, especially in situations where ECLIA are not available, or cannot be reliably used. Diagnostic performances in low prevalence populations still needs to be determined and larger populations need to be tested. In addition, it is clear that serological assay for SARS-CoV-2 antibodies need to be performed at least 10 to 15 days after symptoms onset^7,8^. Finally, here we used plasma and the test was performed in a laboratory environment; we may expect different results in real-life at patients’ bed and using capillary blood.

The second notable finding of this study lies in the fact that IgG and IgM seropositivity is still present 180 days after symptoms onset even with normal antibody decline^9^. To our knowledge, it is the first time that SARS-CoV-2 IgG and IgM seropositivity is demonstrated with RDT 180 DPS. This study indicates also that SARS-CoV-2 IgG/IgM are present constantly and with enough concentration to be detectable with RDT, from 15 days to at least 180 days post symptoms. This finding applies to the this RDT only and must not be applied to other RDTs currently available. It is also necessary to quantitatively determine the level of IgG/IgM 180 days after symptoms onset to confirm this finding obtained with a qualitative assay.

In addition, the test provided clear results, without indeterminate or invalid results (no Control line). There are several limitations to this study. First, we present here the results of a method evaluation study and not a seroprevalence study. Therefore, the PPV obtained here (based on a 26.5% proportion of cases defined as laboratory confirmed SARS-CoV-2 by RT-PCR) will probably be lower in a low prevalence setting. Nevertheless, our results also showed that when targeting a population 10 days after symptoms onset, sensitivity and NPV reach high diagnostic performances (100%). Although not fully established here, we observed that the RDT results were consistent with the other automated serological test (ECLIA) with similar analytical sensitivity. Another limitation of this validation study lies in its limited sample size leading to broad 95% confidence intervals, requiring confirmation of these data at a larger scale. Finally, our present conclusions only apply to this RDT, and must not be applied to any other RDTs currently available.

In conclusion, this RDT is not meant to replace a SARS-CoV-2 RT-PCR diagnostic test in the first week of the disease, but could be a reliable option for assessing the SARS-CoV-2 serology in moderate to high COVID-19 prevalence settings, especially in situations where automated ECLIA or ELISA are not available, with samples collected at least 10 days after symptoms onset and up to 180 days after the onset of symptoms. Further investigations in low prevalence situations and using capillary blood are necessary.

## Data Availability

Data available in Supplementary Material

