## Supplementary material for "Development and evaluation of a new IgM/IgG rapid diagnostic test for SARS-CoV-2"

### GaDia new COVID RDT Evaluation

| RT-PCR Positive samples | | | | | |
|---|---|---|---|---|---|
| **Sample #** | **Age (gender)** | **DPS** | **IgM** | **IgG** | **ECLIA Total Ig (COI)** |
| 1 | 50 (F) | 190 | pos | pos | NT |
| 2 | 53 (M) | 180 | pos | pos | NT |
| 3 | 63 (F) | 15 | pos | pos | NT |
| 4 | 58 (F) | 14 | pos | pos | NT |
| 5 | 63 (F) | 22 | pos | pos | NT |
| 6 | 35 (M) | 17 | pos | pos | NT |
| 7 | 59 (M) | 19 | pos | pos | NT |
| 8 | 33 (M) | 19 | pos | pos | NT |
| 9 | 65 (F) | 17 | pos | pos | NT |
| 10 | 25 (F) | 20 | pos | pos | NT |
| 11 | 54 (F) | 21 | pos | pos | NT |
| 12 | 62 (M) | 36 | pos | pos | NT |
| 13 | 34 (F) | 11 | pos | pos | NT |
| 14 | 48 (M) | 19 | pos | pos | NT |
| 15 | 24 (F) | 16 | pos | neg | NT |
| 16 | 23 (F) | 30 | pos | pos | NT |
| 17 | 40 (F) | 24 | pos | pos | NT |
| 18 | 50 (M) | 14 | pos | pos | NT |
| 19 | 67 (F) | 27 | neg | pos | NT |
| 20 | 48 (F) | 21 | neg | pos | NT |
| 21 | 22 (F) | 22 | pos | pos | NT |
| 22 | 66 (M) | 18 | pos | pos | NT |
| 23 | 51 (M) | 36 | neg | pos | NT |
| 24 | 50 (M) | 16 | pos | pos | NT |
| 25 | 53 (M) | 39 | pos | pos | 180,9 |
| 26 | 12 (M) | 22 | pos | pos | 169,6 |
| 27 | 8 (M) | 20 | neg | pos | 114,9 |
| 28 | 72 (M) | 14 | pos | pos | 91,45 |
| 29 | 5 (F) | 41 | pos | pos | 76,85 |
| 30 | 54 (M) | 16 | pos | pos | 49,05 |
| 31 | 73 (F) | 17 | pos | pos | 20,16 |
| 32 | 40 (F) | 24 | pos | pos | 10,1 |
| 33 | 62 (F) | 15 | pos | pos | 4,98 |
| 34 | 57 (M) | 26 | pos | pos | 2,06 |
| 35 | 36 (F) | 10 | pos | neg | 1,19 |

NT: not tested

| Control Group (collection date < Jan. 2019) | | | | |
|---|---|---|---|---|
| **Sample #** | **Age** | **Gender** | **IgM** | **IgG** |
| 1 | 28 | m | neg | neg |
| 2 | 68 | m | neg | neg |
| 3 | 48 | f | neg | neg |
| 4 | na | na | neg | neg |
| 5 | na | na | neg | neg |
| 6 | na | na | neg | neg |
| 7 | na | na | neg | neg |
| 8 | na | na | neg | neg |
| 9 | na | na | neg | neg |
| 10 | 19 | F | neg | neg |
| 11 | 51 | f | neg | neg |
| 12 | 43 | m | neg | neg |
| 13 | 26 | f | pos | neg |
| 14 | 8 | m | neg | neg |
| 15 | 48 | f | neg | neg |
| 16 | 7 | f | neg | neg |
| 17 | 11 | f | neg | neg |
| 18 | 8 | m | neg | neg |
| 19 | 5 | m | neg | neg |
| 20 | 6 | m | neg | neg |
| 21 | 11 | f | neg | neg |
| 22 | 12 | f | neg | neg |
| 23 | 18 | m | neg | neg |
| 24 | 15 | f | neg | neg |
| 25 | 43 | f | neg | neg |
| 26 | 9 | f | neg | neg |
| 27 | 37 | f | neg | neg |
| 28 | 41 | m | neg | neg |
| 29 | 47 | f | neg | neg |
| 30 | 13 | m | neg | neg |
| 31 | 52 | f | neg | neg |
| 32 | 6 | f | neg | neg |
| 33 | 18 | f | pos | neg |
| 34 | 3 | m | neg | neg |
| 35 | 2 | m | neg | neg |

| | | | | |
|---|---|---|---|---|
| 36 | 13 | f | neg | neg |
| 37 | 3 | f | neg | neg |
| 38 | 38 | f | neg | neg |
| 39 | na | na | neg | neg |
| 40 | na | na | neg | neg |
| 41 | na | na | neg | neg |
| 42 | 32 | M | neg | neg |
| 43 | 23 | M | neg | neg |
| 44 | 45 | F | neg | neg |
| 45 | 49 | F | neg | neg |
| 46 | 31 | M | neg | neg |
| 47 | 55 | M | pos | neg |
| 48 | 19 | M | neg | neg |
| 49 | 80 | F | neg | neg |
| 50 | 62 | F | neg | neg |
| 51 | 35 | F | neg | neg |
| 52 | 23 | M | neg | neg |
| 53 | 33 | M | neg | neg |
| 54 | 55 | M | neg | neg |
| 55 | 36 | M | neg | neg |
| 56 | 32 | M | neg | neg |
| 57 | 26 | M | neg | neg |
| 58 | 35 | f | neg | neg |
| 59 | na | na | neg | neg |
| 60 | na | na | neg | neg |
| 61 | na | na | neg | neg |
| 62 | 40 | f | neg | neg |
| 63 | 29 | f | neg | neg |
| 64 | 10 | f | neg | neg |
| 65 | 53 | f | pos | neg |
| 66 | 24 | m | neg | neg |
| 67 | 70 | f | pos | neg |
| 68 | na | na | neg | neg |
| 69 | 34 | m | neg | neg |
| 70 | 39 | f | neg | neg |

| | | | | |
|---|---|---|---|---|
| 71 | 43 | m | neg | neg |
| 72 | 20 | M | neg | neg |
| 73 | 44 | M | neg | neg |
| 74 | 55 | M | neg | neg |
| 75 | 25 | F | neg | neg |
| 76 | 58 | M | neg | neg |
| 77 | na | na | neg | neg |
| 78 | 42 | M | neg | neg |
| 79 | 52 | M | neg | neg |
| 80 | 34 | M | pos | neg |
| 81 | 43 | M | neg | neg |
| 82 | 41 | M | neg | neg |
| 83 | 25 | F | neg | neg |
| 84 | na | F | neg | neg |
| 85 | 51 | M | neg | neg |
| 86 | na | F | neg | neg |
| 87 | 27 | M | neg | neg |
| 88 | 43 | M | neg | neg |
| 89 | na | na | neg | neg |
| 90 | 34 | M | neg | neg |
| 91 | 29 | F | neg | neg |
| 92 | 38 | M | neg | neg |
| 93 | 29 | F | neg | neg |
| 94 | 34 | M | neg | neg |
| 95 | 29 | F | neg | neg |
| 96 | 34 | M | pos | neg |
| 97 | 31 | M | neg | neg |

na: unknown
